# Delay of TB patients in diagnosis in a conflict setting of Mogadishu, Somalia - a cross-sectional study

**DOI:** 10.1101/2021.11.24.21266678

**Authors:** Naima Said Sheikh, Abdulwahab M. Salad, Abdi A. Gele

## Abstract

**Background:** The TB case detection rate in Somalia is 42%, which is much lower than the WHO target of detecting 70% of new TB cases. Understanding the factors contributing to the delay of TB patients in the diagnosis, and reducing the time between the onset of TB symptoms to diagnosis, is a prerequisite to increase the case detection rate and to ultimately bring the TB epidemic in Somalia under control. The aim of this study is to examine the duration of delay, and factors associated with the delay among patients in TB management centers in Mogadishu, Somalia.

**Methods:** An institution-based, cross-sectional study was conducted in TB management clinics providing directly observed treatments (DOTS) programs in Mogadishu. A total of 276 patients were interviewed using a structured questionnaire from June-October 2018. We analyzed data using descriptive statistics and different logistic regression models.

**Results:** Approximately 78% of study participants were male. Nearly a third (36.5%) came from a household of nine individuals or more, while 73% were unemployed. The median patient and provider delays were 50 days and one day, respectively. The median total delay was 55 days, with an inter-quartile range of 119 days. Patients who had a poor knowledge of the symptoms of TB had 3.16 times higher odds of delay over 50 days than their counterparts. Furthermore, a poor knowledge of the symptoms of TB (aOR 4.22, CI 2.13-8.40), not making one’s own decisions in seeking TB treatment (aOR 2.43, CI 1.22-4.86) and a poor understanding of the fact that TB can be treated with biomedical treatment, as opposed to traditional treatment (aOR 2.07, CI 1.02-4.16), were predictors of a patient delay over 120 days.

**Conclusions:** The duration in the delay of TB patients under diagnosis in Mogadishu is one of the highest reported in developing countries, exceeding two years in some patients. Training local community health workers to detect suspected TB cases, and referring the cases of prolonged cough over three weeks for TB care centers for diagnosis, is imperative to help break the transmission and reduce the infectious pool in the population of Mogadishu. This may not only increase the community awareness of TB disease, but it may also facilitate the early referral of TB patients to diagnostic and treatment care centers.

## Introduction

An estimated one-third of the world’s populations are infected with TB, while an estimated 10.4 million new TB cases and 1.8 million TB-related deaths have occurred in 2015 (1). Despite global progress in tuberculosis control, TB remains a major health burden in fragile states such as Somalia (2, 3). This is attributable to a number of factors, one of them being the large number of infectious TB patients who remain undiagnosed and thus untreated, thereby maintaining the cycle of TB transmission in the community (4). The proportion of cases detected and cured under a directly observed treatment strategy (DOTS) is a reliable indicator that measures the progress, as well as the setbacks of the TB control programs. But the ability of TB programs to increase the cases detected and treated is constrained by factors that deter TB patients from seeking a prompt diagnosis from health-care centers (5).

Somalia is a country experiencing over 3 decades of civil war. The armed conflict have long been linked to the increased incidence of TB through increased population displacement, poverty, and malnutrition (6). The conflict often affect the ability of TB patients to seek prompt care (7). Conflict conditions are also linked to destruction of health infrastructures and interrupted access to anti-TB treatment (6), as well as the subsequent rise in drug resistance TB and exportation of drug-resistant strains to distant countries (8). A previous WHO report estimated for all forms of TB cases in Somalia as 532 per 100,000 persons, and it is apparent that the lack of data has made regular estimates complicated (2). Further, the national TB positive case detection is 32%, and the country has one of the highest multi-drug resistant TB in Africa and the Arab world (3). The Somalia’s TB control programs relies on passive case detection, which means diagnosing infectious cases who present themselves to the health facilities. The passive detection method is dependent on patient’s motivation and knowledge, financial ability, and the availability and effectiveness of diagnostic services. Many parts of the country are still under active armed conflict, and in such areas, substantial number of undiagnosed and untreated infectious cases are likely to remain within the community (6).

While the duration that infectious persons remain within the community untreated is of critical importance in TB control, there is a limited amount of evidence on the duration of delay in the diagnosis of TB patients in Somalia. This study evaluates the delays of TB patients in the diagnosis and factors associated with the delay among the newly diagnosed TB patients in Mogadishu, Somalia.

## Methods

### Study design and participants

An institutional based, cross-sectional study was carried out in Mogadishu, Somalia between June and October 2018. Mogadishu is administratively structured into 17 districts with a total population of 2.4 million. There were nine TB management clinics that provided the DOTS program at the time of the study, and we included all of them in the study. All new TB patients, aged 15 years and above, who were diagnosed clinically or microbiologically within three months from the start of the data collection and consented to participation in the study, were included. Moreover, other patients who were diagnosed during the data collection were also included. A total of 276 TB patients were interviewed from all TB clinics in Mogadishu for the five months of the overall data collection. The study received an ethical approval from the Somali National University’s ethical committee

### Instrument

A structured questionnaire was used for the data collection. Information about socio-demographics, patients’ knowledge about TB, health-seeking behavior and patients’ autonomy to seek care was collected. Patients were asked about major symptoms of TB (persistent cough over three weeks, hemoptysis, fever, chest pain, weight loss, swollen lymph anodes), duration of major presenting symptoms, date of first visit to a professional health-care provider and date of diagnosis. We also asked if they know how TB can be treated using a multiple-choice question involving different traditional remedies used in Somalia and biomedical TB treatment found in the health-care facilities. We also asked if they have spent money for seeking TB care. Initially, an English language questionnaire was developed, then translated to the Somali language, and again back translated to English to ensure the specificity of the questions. The final Somali version was pretested to five new TB patients attending under an intensive phase of TB treatment in the DOTs program. The necessary adjustment was made, and the final version of the questionnaire was prepared. Furthermore, the crosschecking of outpatient cards, patient registration books and laboratory registration forms were made. Two days of training were given for data collectors regarding the content of the questionnaire, and the techniques required for an optimal interview of TB patients.

### Statistical analysis

A statistical analysis was carried out using SPSS version 26, and the data were descriptively summarized. As the data was not normally distributed, nonparametric Chi-square tests (crosstabs) were employed in calculating group differences. To adjust the confounding effect of several identified predictors of diagnostic delay, and to finally establish factors that may independently be associated with the delay, Univariate and multivariate logistic regression analyses was performed. We reported Odds ratio and 95% confidence intervals (95% CI) to establish the association of the variables of interest. Statistical significance was considered at p < 0.05.

### Definitions

*Patient delay*: is defined as the time between the onset of clinical symptoms of TB to the first visit to a professional health-care provider.

*Provider delay*: is defined as the time from a patient’s first consultation with a professional health-care provider for the symptoms of TB until the date of diagnosis.

*Total delay:* is the sum of the patient delay and the medical provider’s delay.

## Results

Table 1 shows the summary of descriptive statistics. A total of 276 participants with a mean age of 30 years (SD ±13.6) were included in this study. Of them, 77.9% were male, 44.9% were married and 48% had no formal schooling. Nearly one-third (36.5%) came from a household of nine individuals or more, while nearly three-fourths (73%) were unemployed. The majority (83%) had pulmonary TB. Of the 221 patients with pulmonary TB, 72% were smear-positive. A larger proportion (84%) had knowledge of the causes of TB, whereas 56% did not know the symptoms of the disease. A vast majority (67%) did not know that TB can be treated with biomedical treatment, and 70% (191) had no knowledge that TB treatment was free of charge. Approximately 37% of the patients consulted a traditional provider prior to seeking biomedical care.

**Table 1:** Characteristics of the study participants.

| Characteristics | Level | N (%) | Characteristics | Level | N (%) |
| --- | --- | --- | --- | --- | --- |
| Age | Mean ± SD; 30.2 ± 13.7 | | | | |
| Gender | Male | 215 (77.9) | TB curable | Yes | 267 (96.7) |
|  | Female | 61 (22.1) |  | No | 9 (3.3) |
| Education | ≥ Secondary | 61 (22.1) | TB medicine free | Yes | 82 (29.7) |
|  | No schooling | 132 (47.8) |  | No | 193 (70.0) |
|  | 1-4 years | 51 (18.5) | Take own decision | Yes | 165 (59.8) |
|  | 5-8 years | 32 (11.6) |  | No | 111 (40.2) |
| Marital Status | Married | 123 (44.6) | Suspect TB | Yes | 151 (54.7) |
|  | Single | 105 (38.0) |  | No | 125 (45.3) |
|  | Divorced/<br>widowed | 46 (16.7) | Tried self-treatment | Yes | 105 (38.0) |
| Occupation | Employed | 75 (27.2) |  | No | 171 (62.0) |
|  | Unemployed | 198 (71.7) | Consulted 1<sup>st</sup> | Public Hosp. | 78 (28.4) |
| Family size | 1 - 4 | 66 (23.9) |  | Private Hosp. | 95 (34.5) |
|  | 5 - 8 | 108 (39.1) |  | Tradition | 102 (37.1) |
| | ≥ 9 | 100 (36.2) | Spent money on TB | Yes | 104 (38.1) |
| Cause Knowledge | Good | 227 (82.2) |  | No | 169 (61.9) |
|  | Poor | 46 (16.7) | Health-care provider 1<sup>st</sup> visited | Private | 109 (39.6) |
| Symptom knowledge | Good | 120 (43.5) |  | Public | 166 (60.4) |
|  | Poor | 155 (56.2) | Treatment | Failure, relapse, default | 15 (5.5) |
| Transmission knowledge | Good | 209 (75.7) |  | New | 256 (94.5) |
|  | Poor | 66 (24.0) | TB type | Pulmonary | 225 (83.3) |
| Diagnosis knowledge | Good | 141 (51.1) |  | Extra pulmonary | 45 (16.7) |
| | Poor | 135 (48.9) | | $\leq 50$ | 138 (50.2) |
| Treatment knowledge | Good | 89 (32.2) | Patient delay | >50 | 137 (49.8) |
|  | Poor | 185 (67.2) |  |  |  |

### Delays

The study found a median patient delay of 50 days, a median provider delay of one day, and a median total delay of 55 days. As shown in Table 2, a median patient delay of 50 days and a mean delay of 81 were found (the 25^th^ and 75^th^ percentile were 14 and 135, respectively). A significant difference was found between those who had a good knowledge of the symptoms of TB and those who did not (p=0.000). Approximately 61% of those who had a poor knowledge about the symptoms of TB had a median delay >50 days, with a majority having a delay over 120 days. Similarly, a significant difference was found between those who visited a public health-care facility and those who visited a private health-care facility (p=0.001).

**Table 2:** Distribution of delays (days)

| Delay (days) | Median | 25<sup>th</sup> percentile | 75<sup>th</sup> percentile | Inter-quartile range (IQR) |
| --- | --- | --- | --- | --- |
| Patient delay | 50 | 14 | 135 | 121 |
| Health-care provider delay | 1 | 0 | 2 | 2 |
| Total delay | 55 | 20 | 139 | 119 |

### Patient delay

There was no significant difference with regard to patient delay by gender, education, age, marital status and occupation. A multivariate logistic regression analysis showed that a poor knowledge of TB symptoms (aOR. 3.16, CI 1.82-5.49), type of health-care provider visited first (aOR 0.31, CI 0.14-0.68), and having spent money on TB diagnosis and treatment (aOR 2.26, CI 1.05-4.87), were predictors for patient delay >50 days (Table 3). However, 29% of participants had a delay of over 120 days. A poor knowledge of the symptoms of TB (aOR 4.22, CI 2.13-8.40), not making one’s own decisions in seeking TB treatment (aOR 2.43, CI 1.22-4.86) and a poor understanding of the fact that TB can be treated with biomedical treatment, as opposed to traditional treatment (aOR 2.07, CI 1.02-4.16), were all predictors of a patient delay over 120 days.

**Table 3:** Factors associated with patient delays.

| Variables | Patient delay<br>≤50 days | Patient delay<br>>50 days | Crude | Adjusted |
| --- | --- | --- | --- | --- |
| **Gender** |  |  |  |  |
| Male | 105 | 109 | ref | ref |
| Female | 33 | 28 | 1.22 (0.69-2.16) | 0.95 (0.48-1.91) |
| **Age** |  |  |  |  |
| ≤20 | 41 | 37 | ref |  |
| 21-51 | 82 | 89 | 0.83 (0.48-1.42) | 1.01 (0.6-2.24) |
| 52+ | 15 | 11 | 1.23 (0.50-3.01) | 1.63 (0.49-5.46) |
| **Marital status** |  |  |  |  |
| Married | 57 | 66 | ref |  |
| Divorced/widowed | 25 | 21 | 1.30 (0.77-2.19) | 1.81 (0.82-3.95) |
| Single | 55 | 49 | 1.37 (0.69-2.72) | 1.60 (0.73-3.51) |
| **Occupation** |  |  |  |  |
| Employed | 28 | 47 | ref | ref |
| Unemployed | 109 | 89 | 2.05 (1.19-3.54) | 1.83 (0.98-3.41) |
| **Education** |  |  |  |  |
| Secondary or above | 34 | 26 | ref | ref |
| 5-8 years | 18 | 14 | 0.98 (0.41-2.33) | 1.40 (0.52-3.77) |
| 1-4 years | 20 | 31 | 0.49 (0.23-1.05) | 0.91 (0.42-1.96) |
| No formal education | 66 | 66 | 0.76 (0.41-4.88) | 0.96 (0.28-1.68) |
| **Spent money on the diagnosis and treatment of TB** |  |  |  |  |
| No | 88 | 81 | ref |  |
| Yes | 50 | 53 | 0.86 (0.53-1.41) | 2.26 (1.05-4.87)* |
| **Symptoms knowledge** |  |  |  |  |
| Good | 78 | 42 | ref |  |
| Poor | 60 | 94 | 2.80 (1.62-4.83)* | 3.16 (1.82-5.49)* |
| **Knowledge of biomedical treatment** |  |  |  |  |
| Good | 51 | 38 | ref |  |
| Poor | 85 | 99 | 0.64 (0.38-1.06) | 0.76 (0.43-1.35) |
| **Type of health-care provider visited first** |  |  |  |  |
| Private | 63 | 45 | ref |  |
| Public | 74 | 92 | 1.77 (1.8-2.90) * | 0.31 (0.14-0.68)* |
| **Did you take decision of seeking treatment** |  |  |  |  |
| Yes | 86 | 78 | ref | ref |
| No | 52 | 59 | 0.80 (0.49-1.29) | 0.66 (0.37-1.19) |

### Health provider delay

The median health provider delay was ‘1’ with mean of ‘5’. Approximately 35% of study participants had a health provider delay over two days. Almost 50% of those who used self-treatment for their illness prior to seeking health care had two days or more of health provider delays than their counterparts (p=0.02). A multivariate logistic regression analysis showed that a poor knowledge on the biomedical treatment of TB versus traditional treatment is the only predictor for a health provider delay longer than two days (aOR 1.76, CI 1.04-3.01).

### Total delay

We found a median total delay of 55 days, with a mean of 86 days. The highest delay was 899 days. However, approximately 40% of participants had a total delay that exceeded three months (90 days).

## Discussion

This study is the second of its kind to report a diagnostic delay of TB patients in Somalia, and reveals an extremely long delay in the diagnosis of TB. Identifying the length of delays, where delays occur and the reasons for the delay will help TB control programs in Somalia improve their control strategies. A median patient delay of 50 days, with mean of 81 days, was found. These figures are almost similar, with the median and mean patient delay (53 and 77) found by a previous study in Somalia (9). Even so, 29% of patients in our study had a delay of over 120 days. This a very serious concern, given the fact that such a long delay leads to continued transmission, which increases the chances of complications and mortality due to TB (10).

Factors associated with patient delay in Mogadishu include a poor understanding of TB symptoms among TB patients. Generally speaking, Somalis have a low health literacy (11). In its early stage, tuberculosis presents with non-specific symptoms that can be easily confused with other illnesses. Thus, the disease may progress for months before patients suspect that they have TB. Nonetheless, if patients are not familiar with the typical symptoms of TB, he\she may barely seek biomedical care (4). There is established evidence about the association between a poor knowledge of TB and a long patient delay (12). Patients with a poor understanding of TB symptoms may feel easy, with non-incapacitating early symptoms of TB that are sometimes compatible with day-to-day activities until they are severe enough to warrant concern. Particularly in Somalia, where due to prevalent poverty and insecurity people struggle with daily lives, it is unlikely for the breadwinner of a family to seek care for symptoms that are not severe enough. A patient who opts to work for the family, and delays seeking care until his\her situation deteriorates, is called “*bukaan socod”* in Somalia, which translates to *walking sick*, and this decision is often considered as a sign of strength. Reducing a delay in the diagnosis of TB patients in Somalia is possible, provided that we fully understand the cultural and systemic factors behind the delay, and accordingly develop and implement an evidence-based intervention that is tailored to local conditions. For example, in Rwanda, the median diagnostic delay of TB patients decreased from 88 days to one day, and a treatment delay from 76 days to three days within a period of 10 years (13). This drastic reduction of the delay in diagnosis was achieved through the implementation of evidence-based interventions that are economically feasible, culturally acceptable and can be applied using locally available resources. The delay in the diagnosis of TB patients in Mogadishu, where most of the diagnostic and treatment facilities in the country are concentrated, is extremely high; hence, the logic supports that a delay in the diagnosis of TB patients in rural areas in Somalia may be alarmingly higher. Data shows that people in rural areas have a significantly higher patient delay than those in urban areas in low and middle income countries (14). In Mogadishu and Somalia, where the literacy rate is approximately 20%, and given that health literacy is low among Somalis (11), an increased awareness about the symptoms of TB, and the deadly consequences of TB if not treated early enough should be considered among people in Mogadishu.

Not making one’s own decision in seeking a TB diagnosis and treatment was associated with a patient delay >120 days. In the Somali community, the decision of seeking care often goes through multiple layers of advisory discussions involving parents, relatives, neighbors and people with an experience of similar illnesses (4). A prior study reported a similar finding, in which the patient’s decision to seek TB treatment is influenced by family and relatives (15). Sometimes, biomedical treatment or traditional remedies are borrowed from former patients who have experienced resampling symptoms, and who believed that the given remedy helped cure their symptoms. A previous study found that seeking biomedical TB care is often the final option for many Somali families, in part because of direct and indirect costs associated with TB treatment (4). It is worth noting that 70% of our study participants did not know that TB treatment was free of charge. This finding is in accordance with prior findings that costs incurred by TB patients lead to a delay of TB patients in seeking a prompt diagnosis (16), which is further confirmed by our findings that patients who spent money on seeking TB diagnosis and treatment had an over twofold higher odds of delay than those who did not spend any money.

A poor knowledge of the fact that TB can be treated with biomedical treatment, as opposed to traditional remedies, was a predictor for both a long patient delay over 120 days and a health provider delay over two days. Given the prolonged civil war, a deprivation of quality health care among Somalis in Somalia may compel people to rely more on the traditional system. It is worth noting that traditional treatment and religious healing are often available free of charge or extremely cheap. A prior study reported that Somali pastoralists seek traditional treatment for TB, and if symptoms do not subside, religious remedies are applied in the form of Koranic verses read for the patient as a second option (4). Only then do they seek modern medicine as an alternative when all other available traditional options have failed, and patients have reached a critical stage of illness. As this may delay patients from seeking biomedical care, a study shows that even if they seek and partly trust biomedical treatment, they still believe that it must be complimented by other practices, which may delay their initiation of TB treatment (4). The health-care system in Somalia is weak, geographically inaccessible to the most vulnerable and too expensive for the majority, which might affect people’s trust toward biomedical treatment, in contrast to traditional treatment that is often readily available in the vicinity, and is often free of charge. The Ministry of Health and non-governmental organizations (NGOs) that provide TB services to people in Mogadishu and Somalia should generally turn their focus to an awareness of TB symptoms, TB treatment and the importance of seeking early diagnosis and treatment.

This study has its limitations. Patients’ inability to recall the dates and entire pathway of the onset of TB symptoms could possibly lead to the collection of suboptimal information. Additionally, low levels of health literacy and a poor knowledge of TB by patients may have thwarted the gathering of the required information. However, experienced researchers conducted the interviews using the local calendar, major national days and festivals to determine patients’ perceived date of onset of TB symptoms. Despite these limitations, the present study is only the second of its kind to examine delays in the diagnosis of TB conducted in Mogadishu for the last 30 years of the civil war. Therefore, the study provides context-specific evidence that we believe will contribute to informing efforts to control TB in the country.

## Conclusion

The study found a long delay in the diagnosis (median of 50 days) of TB patients in Mogadishu. This being the capital city, worse situations may be expected in other parts of the country with much less access to TB diagnostic and treatment services. The results indicate patients who do not make their own decisions, patients with a poor knowledge of TB symptoms, and those who have a poor knowledge of the biomedical treatment of TB, have longer patient and health provider delays than their counterparts. Training local community health workers to detect suspected TB cases and refer them to TB management centers for diagnosis and treatment is imperative to help break the transmission and reduce the infectious pool in the population in Mogadishu. This may not only increase community awareness of TB, but may also facilitate an early referral of TB patients to diagnostic and treatment care centers.

## Data Availability

All data produced in the present work are contained in the manuscript

## Acknowledgments

We would like to thank Galad Dahir, Mohamed Hussein and Liban Gaylac for collecting the data.

## Ethics Statement

Ethical approval was received from Somali National University. Informed consent was obtained from each participant prior to the procedure.

